# A Droplet Digital PCR Assay to Detect SARS-CoV-2 RNA

**DOI:** 10.1101/2020.05.06.20090449

**Authors:** Martin J. Romeo, Christian P. DiPaola, Cassidy Mentus, Cynthia D. Timmers

## Abstract

We describe a quantitative droplet digital PCR (ddPCR) assay for detection of SARS-CoV-2 viral ribonucleic acid (RNA) in total RNA extracted from human sputum. This method was validated using the guidance of the United States Food and Drug Administration’s Accelerated Emergency Use Authorization (EUA) Template for SARS-CoV-2 that Causes Coronavirus Disease (COVID-19) Molecular Testing of Respiratory Speciment in CLIA Certified High-Complexity Laboratories. Though our laboratory is not CLIA certified, this method met all criteria specified by the guidance document with a Limit of Detection (LOD) of 0.25 copies/μL in the final ddPCR (at least 19/20 replicates reactive), which we consider to be a Lower Limit of Quantification (LLOQ); inclusivity of all known annotated SARS-CoV-2 genomes; no cross-reactivity with other respiratory pathogens; and reactivity of all contrived positives at or above the LOD.

## Introduction

Our laboratory has developed and validated a droplet digital polymerase chain reaction^1,2^ (ddPCR) method to detect SARS-CoV-2 (novel coronavirus/2019-nCoV) nucleic acids in human sputum specimens that is more than an order of magnitude more sensitive than real-time reverse-transcriptase polymerase chain reaction (rRT-PCR)^3^, which is the current gold standard for clinical evaluation of active SARS-CoV-2 infection. This method is a quantitative endpoint assay in which RNA extracted from human sputum is reverse transcribed before the resulting cDNA is added as template to a PCR mix. The 20 μL PCR is then partitioned into ~20,000 1-nL droplets in an oil emulsion before target sequences are amplified, with each droplet that passes a size filter being analyzed for fluorescence intensity in two channels after 40 cycles of amplification. After reading the droplets, thresholds are applied to the fluorescence intensities of those two channels to discriminate between positive (target amplified) and negative (no target amplified) droplets, and the resulting concentration of target sequence, in copies/μL, can be calculated based on the number of positive and negative droplets in each reaction. Partitioning is critical for detection of low-abundance targets and reduces the ability of inhibitors to negatively impact amplification^4^. This methodology enables far more sensitive detection^5^ of SARS-CoV-2, particularly for early diagnosis in infected individuals who will become symptomatic and for asymptomatic individuals with low viral loads that may be missed by rRT-PCR tests.

The QX200 ddPCR system (Bio-Rad, Hercules, CA) has been, in our experience, a reliable and robust platform for automated droplet generation and droplet reading (autoDG and QX200, respectively). The method we describe here utilizes commercially available reagents and kits for both reverse transcription and ddPCR. Automated RNA extraction from human sputum samples was performed with simplyRNA from Tissue kits in a Maxwell® RSC (both from Promega, Madison, WI). Complementary DNA (cDNA) was reverse transcribed from RNA extracts using SuperScript™ IV VILO™ Master Mix (Invitrogen by Thermo Fisher Scientific, Carlsbad, CA). The PCR primer/probe sequences for both SARS-CoV-2 (N1 region of the nucleocapsid gene) and human RNase P/RPP30 (RP) have been previously published by the United States Department of Health and Human Services^6^; the N1 probe is conjugated to FAM reporter and BHQ-1 quencher, whereas the RP probe is conjugated to HEX reporter and BHQ-1 quencher (Integrated DNA Technologies, Coralville, IA). Purified SARS-CoV-2 RNA was generously supplied by the UTMB World Reference Center for Emerging Viruses and Arboviruses (Galveston, TX).

## Methods

Briefly, sputum was collected from laboratory members in 1.5 mL DNA LoBind tubes (Eppendorf) and placed on ice. 200 μL of Homogenization Buffer containing 1-thioglycerol (prepared per user manual for the simplyRNA from Tissue kit) was added to each specimen; the tubes were capped securely and vortexed vigorously for 30 seconds. Lysis Buffer (200 μL) was added to each homogenate and the capped tubes were again vortexed vigorously for 30 seconds. The entire volume of lysate was transferred with a wide-bore pipette tip to the simplyRNA cartridges, which were prepared as described in the user manual and processed with the Maxwell RSC according to the simplyRNA Tissue method uploaded to the instrument. Purified RNA was eluted in 50 μL of nuclease-free water (water and 0.6 mL elution tubes provided in the kit). Typical RNA yield was in the 25-500 ng/μL range. Spike-in experiments for the creation of "contrived" (synthetic) positives was performed with purified SARS-CoV-2 RNA from an appropriate substock, as we empirically determined the concentration of the 100 ng/μL stock provided by UTMB WRCEVA to be ~8.0 × 10^8^ copies/μL. We performed serial 10-fold dilutions in low TE (10 mM Tris, 0.1 mM EDTA) buffer and added purified viral RNA to non-reactive RNA extracts such that the spike volume did not exceed 10% of the total final volume of the contrived positive samples. Naked, purified viral RNA added to human sputum before extraction, even in the presence of 1-thioglycerol and kept on ice, could not be detectably recovered presumably due to rapid degradation by RNases in the specimens.

Reverse transcription was required to convert RNA to cDNA, which is a suitable template for amplification by DNA polymerase in the PCR^3^. The cDNA synthesis reactions were carried out in 8-strip PCR tubes to facilitate transfer of cDNA to the ddPCR “donor” plate. A RT reaction mix consisting of SuperScript™ IV VILO™ master mix (4.4 μL per reaction) and nuclease-free water (12.1 μL per reaction) was mixed by brief vortexing and then centrifugation to collect the liquid at the bottom of the reaction mix tube, then maintained on ice. 15.0 μL of RT reaction mix was added to each strip tube needed for the assay, followed by 5.0 μL of the appropriate RNA extract or PC (Positive Control; non-reactive human sputum RNA extract containing 400 copies/μL purified SARS-CoV-2 RNA). RT reactions containing RNA were mixed by gently pipetting up and down 15 times with a multichannel pipette set to 15.0 μL. The strip tubes were centrifuged briefly to collect liquid at the bottom of the tubes and placed in a C1000 thermal cycler (Bio-Rad, Hercules, CA) with the following parameters (heated lid set to 85°C with a sample volume of 20 μL):

| **<u>Cycling Step</u>** | **<u>Temperature (°C)</u>** | **<u>Time</u>** | **<u>Number of Cycles</u>** |
| --- | --- | --- | --- |
| Primer annealing | 25 | 10 min | 1 |
| Reverse Transcription | 53 | 10 min | 1 |
| Enzyme inactivation | 85 | 1 min | 1 |
| Hold | 4 | infinite | 1 |

A ddPCR reaction mix consisting of ddPCR Multiplex Supermix (5.5 μL per reaction), N1 primer/probe mix (2.75 μL per reaction), RP primer/probe mix (2.75 μL per reaction) and nuclease-free water (5.5 μL per reaction) was prepared by vortexing briefly, then centrifuged to collect the liquid at the bottom of the tube (final concentration of each primer was 825 nM and each probe was 230 nM). The ddPCR reaction mix was then maintained on ice until cDNA synthesis was completed. From this point on, ddPCR plate set-up was performed in a tissue culture hood, on ice, to minimize dust contamination. 15.0 μL of the ddPCR reaction mix was added to the appropriate wells of a fresh 96-well ddPCR plate, and 5.0 μL of cDNA was added to the wells designated on a plate map. NC (negative control), in this case a true NTC, or No Template Control, is 5.0 μL nuclease-free water added to the ddPCR reaction mix. **20 μL of Buffer Control for Probes, diluted to 1X, was added to the empty wells of any plate columns containing ddPCR reactions**. Once samples, controls and Buffer Control were added to the 96-well ddPCR plate, the final reactions were mixed by gently pipetting up and down 15 times with a multichannel pipette set to 15.0 μL. A plate seal was loosely placed over the 96-well ddPCR plate to prevent entry of particulates into wells containing liquid. The ddPCR plate (“donor plate”) was sealed using a PX100 plate sealer (Bio-Rad, Hercules, CA) and immediately placed on ice again to prevent initiation of the reaction. After sealing, plates were centrifuged at 1000 × *g* for 30 seconds at ambient temperature to collect the liquid in the bottoms of the plate wells and maintained on ice during droplet generator setup.

The droplet generator (autoDG) was configured by selecting all columns containing samples, controls or Buffer Control, and naming the run (for example, “SARS-CoV-2 sample analysis 23Apr2020 MJR”) and adding notes as needed. Droplet Generation Oil for Probes was used. Sufficient fresh, unused cartridges were loaded to accommodate the number of plate columns selected during configuration, and enough filter tips were present such that for each column on the sample plate there were TWO columns of tips. The sealed donor plate was placed carefully but securely in the clips on the donor plate platform inside the autoDG. A cold chill block (stored at −20°C, purple side down) was placed in the autoDG with the “cyclops” eye on the bottom and toward the back of the autoDG using the alignment stubs located on the platform. A fresh, unused 96-well ddPCR plate (“acceptor plate”) was placed securely on the cold chill block in the autoDG in the same orientation as the donor plate. Droplet generation was initiated after ensuring all components were present in sufficient quantities for the run. After droplet generation was completed, the acceptor plate was sealed with a fresh, unused plate seal and visually inspected to ensure uniformity of the milky emulsion layer across all wells containing samples/controls/Buffer Controls. After generation, droplets were particularly fragile and care was taken to handle the sealed acceptor plate gently; thermal cycling was initiated within 45 minutes of completing droplet generation as recommended by the manufacturer. The sealed acceptor plate was then placed in a C1000 thermal cycler with the following parameters (heated lid set to 105°C with a sample volume of 40 μL):

| <u>Cycling Step</u> | <u>Temperature (°C)</u> | <u>Time</u> | <u>Ramp Rate</u> | <u>Number of Cycles</u> |
| --- | --- | --- | --- | --- |
| Enzyme Activation | 95 | 10 min |  | 1 |
| Denaturation | 94 | 30 sec | 2°C/sec<br>for all | 40 |
| Annealing/extension | 59 | 1 min |  | 40 |
| Enzyme deactivation | 98 | 10 min |  | 1 |
| Hold | 4 | infinite |  | 1 |

Once the thermal cycler reached 4°C, the thermal cycling protocol was canceled. The ddPCR plate was then allowed to equilibrate to ambient temperature for 5 minutes. The droplet reader as turned on and the QuantaSoft program was opened on the computer linked to the droplet reader. NOTE: NO PRIMING, FLUSHES OR DECONTAMINATION WERE REQUIRED. However, if the droplet reader was inactive for more than 3 days, a prime was performed to maintain consistent droplet reading results, especially in well A1 of the ddPCR plate. The droplet reader plate chamber was opened, the ddPCR plate was loaded and the chamber was closed. The droplet reader template was configured according to the PLATE MAP used for donor plate set up (double click in at least one cell, e.g. A1). Sample identifiers were added for all wells containing samples or controls (for example, “S1”, “NTC”, “PC”). Under the “EXPERIMENT” dropdown menu, “ABS” was selected. In the “SUPERMIX” dropdown menu, “ddPCR Supermix for probes <no dUTP>” was selected. For Target 1, “N1” was entered in the "Name” box and the appropriate sample type was selected (“Ch1 Unknown”, “Ch1 Positive”, “NTC” as appropriate). For Target 2, “RP” was entered in the “Name” box and the appropriate sample type was selected (“Ch2 Unknown”, “Ch2 Positive”, “NTC” as appropriate). “Apply” (blue text) must be clicked to save those selections in the desired wells. After selecting “OK” (blue text), droplet reading was initiated by clicking the “Run” icon to the left of the plate template, then selecting the “FAM/HEX” dye set in the follow-up window. Data were analyzed manually by applying a threshold of 6000 (Amplitude) to all N1 wells and 4000 (Amplitude) to all RP wells.

## Results

By applying this method to contrived positive RNA extracts, we were able to achieve what we consider a Lower Limit of Quantification/LLOQ (though defined as LOD in the EUA guidance document) of 0.25 copies/μL of viral N1 in the final ddPCR, or 4 copies/μL of viral RNA in the total RNA extract (77.7% accuracy, 19.4% CV) with 19/20 individually spiked replicates demonstrating reactivity. The true LOD for this method is quite likely below 0.25 copies/μL as we were able to detect reactivity in 2 of 2 contrived positives spiked at 2 copies/μL in the RNA extract, but unable to detect reactivity in a contrived positive spiked at 1 copy/μL. All contrived positive samples containing at least 4 copies/μL of purified viral RNA in the total RNA extract demonstrated reactivity in a clinical evaluation analysis (at least 20 contrived positives at 1x-2x LOD as defined by EUA guidance), with 2 of 3 contrived positives below LLOQ yielding positive droplets. *In silico* analysis of the N1 and RP primer/probe sequences, along with wet testing of plasmid-borne SARS-CoV and MERS-CoV nucleocapsid gene sequences during method development, reveal 100% inclusivity of known annotated SAS-CoV-2 genomes and no cross-reactivity of common and related respiratory pathogens. We consider this method validated in accordance with the criteria set forth in the United States Food and Drug Administration’s ACCELERATED EMERGENCY USE AUTHORIZATION (EUA) TEMPLATE FOR SARS-COV-2 THAT CAUSES CORONAVIRUS DISEASE (COVID-19) MOLECULAR TESTING OF RESPIRATORY SPECIMENS IN CLIA CERTIFIED HIGH-COMPLEXITY LABORATORIES^7^, with the provision that our laboratory is not CLIA-certified.

### Instruments

A. Bio-Rad PX1 PCR Plate Sealer (181-4000)
B. Bio-Rad C1000 Touch Thermal Cycler with 96-Deep Well Reaction Module (185-1197)
C. Bio-Rad Automated Droplet Generator (autoDG; 186-4101)
D. Bio-Rad QX200 Droplet Reader (186-4003)
E. Computer with QuantaSoft software

### Reagents and Consumables

A. 0.6 mL tubes (Axygen, MCT-060-L-C)
B. 1.5 mL DNA LoBind tubes (Eppendorf, 022431021)
C. TempAssure 0.2mL PCR 8-tube strips (USA Scientific, 1402-4700)
D. ddPCR Multiplex Supermix (Bio-Rad, 12005910)
E. Nuclease-free water (Ambion, AM9937)
F. 96-well semi-skirted deep well plates (Bio-Rad, 12001925)
G. Plate seals (Bio-Rad, 1814040)
H. Droplet Generation Oil for Probes (Bio-Rad, 1864110)
I. DG32 cartridges (Bio-Rad, 1864108)
J. 2x ddPCR Buffer Control for Probes (Bio-Rad, 1863052)
K. SuperScript™ IV VILO™ Master Mix kit (Life Technologies, 11756500)
L. Droplet Reader Oil (Bio-Rad, 1863004)
M. N1 primer/probe set from 2019-nCoV RUO Kit (Integrated DNA Technologies, 10006713)
N. Custom RP primer/probe set (Integrated DNA Technologies)
O. Positive Control: 2019-nCoV_N plasmid (Integrated DNA Technologies, 10006625) Note: Dilute positive control plasmid DNA to 500 copies/μL in nuclease-free water prior to ddPCR.

## Data Availability

All data referred to in the manuscript are available upon request

